# Viral genetic sequencing identifies staff transmission of COVID-19 is important in a community hospital outbreak

**DOI:** 10.1101/2021.02.18.21250737

**Authors:** Jane A.H. Masoli, Aaron Jeffries, Ben Temperton, Cressida Auckland, Michelle Michelsen, Joanna Warwick-Dugdale, Robyn Manley, Audrey Farbos, Sian Ellard, Beatrice Knight, Claire Bewshea, Christine Sambles, James Harrison, Ben Bunce, Alexis Carr, Andrew T. Hattersley, Stephen Ll. Michell, David J. Studholme

**Affiliations:** Epidemiology and Public Health Group, College of Medicine and Health, University of Exeter, Exeter, UK; Royal Devon and Exeter Hospital NHS Trust, Barrack Road, Exeter, UK; Biosciences. College of Life and Environmental Sciences, University of Exeter, Exeter, UK; Institute of Biomedical and Clinical Science, College of Medicine and Health, University of Exeter, Exeter, UK

## Abstract

**Background:** We have successfully used whole-genome sequencing to provide additional information for transmission pathways in infectious spread. We report and interpret genomic sequencing results in clinical context from a large outbreak of COVID-19 with 46 cases across staff and patients in a community hospital in the UK.

**Methods:** Following multiple symptomatic cases within a two-week period, all staff and patients were screened by RT-PCR and staff subsequently had serology tests.

**Results:** Thirty staff (25%) and 16 patients (62%) tested positive for COVID-19. Genomic sequencing data showed significant overlap of viral haplotypes in staff who had overlapping shift patterns. Patient haplotypes were more distinct from each other but had overlap with staff haplotypes.

**Conclusions:** This study includes clinical and genomic epidemiological detail that demonstrates the value of a combined approach. Viral genetic sequencing has identified that staff transmission of COVID-19 was important in this community hospital outbreak.

**Key points:**

- Detailed analysis of a large community hospital outbreak in older adults and staff with concurrent clinical and genomic data, including working patterns.
- Staff transmission was important in this community hospital outbreak.
- We found plausible associations between staff and patient cases.

## Introduction

Whole-genome sequencing and phylogenetic analysis of pathogen isolates can be combined with traditional epidemiology to define transmission pathways and track outbreaks.^1^ Applied to bacterial pathogens such as methicillin-resistant *Staphylococcus aureus* (MRSA),^2^ sequencing has distinguished between closely related genotypes that would be otherwise indistinguishable, thereby improving the resolution of analyses of pathogen transmission. Viruses have high rates of mutation and replication, resulting in rapid genetic variation.^3^ It has been proposed that viral genome sequencing and phylogenetic analyses could be key in viral epidemics by adding molecular precision^4^ and this approach was pioneered in near real time during the West African ebola epidemic of 2014 to 2016^5^. The SARS-CoV-2 pandemic has precipitated unprecedented levels of application of viral genome sequencing, which has been used to inform clinical decision-making.^5^ Clinically, spread of COVID-19 can occur in the community or within hospitals and can be acquired from multiple sources. Outbreak analysis best practice combines detailed clinical epidemiology with supporting microbiological testing.^6^ However, it is extremely difficult to ascertain the source of a positive case and standard testing by reverse transcriptase–polymerase chain reaction (RT-PCR)^7^ does not aid understanding of the transmission pathway. Phylogenetic analysis of genome sequences from outbreaks can add essential detail. This is exemplified in New Zealand where genomic surveillance has been successfully deployed to understand the origins of SARS-CoV-2 in circulation and patterns of their transmission.^8^

The COVID-19 Genomics UK (COG-UK) Consortium^9^ was established to support rapid whole-genome viral sequencing for confirmed COVID-19 cases^10^. As of June 2020, the epidemic in the UK comprised at least 1356 independently introduced lineages. Comparison of the genome sequences of viral isolates from a cluster of cases can be used to test hypothetical epidemiological links between those cases and provide supporting or contradictory evidence.

We report the analysis of a clinical outbreak of COVID-19 at a community hospital in Devon, United Kingdom, and show whole-genome viral sequencing combined with clinical context to hypothesise likely transmission routes in this outbreak.

## Methods

### Study design and participants

We investigated a COVID-19 outbreak that occurred in a 32-bed community hospital with 118 staff. The community hospital focuses on rehabilitation, with the majority of patients transferred following inpatient stays at the acute hospital.

### COVID-19 testing

The full inpatient cohort of 26 and 118 staff were tested for COVID-19 by reverse transcriptase– polymerase chain reaction (RT-PCR) on nasopharyngeal and throat swabs within the outbreak period.

RT-PCR testing used the platforms SmartCycler® (Cepheid, Sunnyvale, CA, USA) and ELITe InGenius® (ELITech Group, Torino, Italy).

Serological testing of COVID-19 via antibody detection was introduced in the UK in May 2020. Antibody testing on serology was performed in 115 staff when first available in the UK NHS, seven weeks after the initial RT-PCR screening. Total antibody count has reported to yield up to 100% sensitivity from two weeks after symptom onset^7,11^. Serology was performed to ascertain whether initial screening at outbreak accounted for the majority of cases in the staff.

### Clinical analysis

Detailed information was obtained for clinical epidemiological evaluation from outbreak surveillance and supported by additional detail for participants recruited to the Exeter COVID-19 Sequencing project (ExCoSe). The sources used for clinical analysis consisted of inpatient medical records, staff symptom questionnaires, and staff rotas. We ascertained symptom timelines, exposures, co-working, COVID-19 severity and outcomes.

### Viral genome sequencing

44 of 46 positive cases had retained swab material from the RT-PCR on which whole-genome viral sequencing was attempted. RNA was prepared and sequenced using protocols published by the ARTIC network (https://artic.network/ncov-2019), based on 400 bp PCR tiling amplicons to sequence the whole 30 kb genome. Briefly, RNA extracted from nasopharyngeal swabs was reverse transcribed and used as a template in for two separate multiplexed PCRs for 35 cycles. The two multiplexed PCRs were then pooled for each sample, purified with 1x AMPure XP bead purification (Beckman Coulter, USA), quantified by fluorimetry and normalized in concentration.

Sequencing libraries were prepared (Oxford Nanopore Technologies, UK) by adding barcode and adaptor to the amplicon DNA; the full protocol is available at https://dx.doi.org/10.17504/protocols.io.bdp7i5rn. Sequencing was performed on an Oxford Nanopore MinION. Each sequencing run comprised 23 samples plus one negative control (carried through from RT-PCR step). Duration of sequencing runs sequenced were 16 to 24 hours, with high-accuracy real-time base calling and demultiplexing (Guppy 3.5.2) and monitoring with MinKNOW v3.6.5 (Oxford Nanopore Technologies) and RAMPART^12^. The ARTIC pipeline (https://github.com/artic-network/artic-ncov2019.git) was then used for subsequent filtering, assembly and variant calling.

### Sequence Analysis

We aligned genome sequences against the Wuhan-Hu-1 reference genome sequence (GenBank MN908947.3^13^) using MAFFT v7.310. ^14^ Haplotypes were inferred from the resulting alignment using a custom script (https://github.com/davidjstudholme/get_viral_haplotypes) that identifies genomic sites that show variation and are unambiguously called (i.e. no Ns) across all analysed genome sequences to generate a Nexus-formatted output file.^15^ This Nexus file served as input into Popart ^16^ to generate median-spanning networks.^17^

### Ethical Approval

The genomic sequencing was performed as part of the COG-UK study. COG-UK genomic sequencing is conducted as part of surveillance for COVID-19 infections under the auspices of Section 251 of the NHS Act 2006. The COG-UK study protocol was approved by the Public Health England Research Ethics Governance Group (reference: R&D NR0195).

Participants were identified during routine clinical practice and recruited via the Royal Devon and Exeter Tissue Bank (RDETB – ethical approval REC no 11/SW/0018) set up to facilitate research through the collection of “spare” clinical samples available at the time of routine procedures.

Approval for the ExCoSe study (STB63/CTB58) was obtained from the RDETB Steering Committee, who are delegated to approve study specific collections. The RDETB is managed within the NIHR Exeter Clinical Research Facility.

## Results

### Description of outbreak

Multiple cases were detected, following the onset of symptoms in both staff and patient groups, therefore screening was conducted on all inpatients (n=26) and staff (n=118). **PCR Testing** RT-PCR tested positive in 30 of 118 staff (25%) and 16 of 26 (62%) patients.

### Staff cases

Six of the staff cases were asymptomatic at the point of testing, with two remaining asymptomatic. The majority developed mild to moderate symptoms at worst but one case had severe COVID-19 requiring hospitalisation. The first cases of symptomatic staff members preceded any positive results in inpatients. All symptomatic staff remained away from work appropriately. This outbreak occurred before routine testing was available for healthcare workers. Two positive staff members had been symptomatic, followed Public Health England guidance on absence and return to work and had returned to work following their period of absence prior to their positive COVID-19 swab.

### Patients

The mean age of the 16 COVID-19 positive patients was 82 years (SD 8.7) and 63% were female. All of the COVID-19 positive patients had recently been inpatients at the acute hospital trust prior to transfer to community hospital, with a mean time from transfer to confirmed COVID-19 of 18 days (SD 9.5). Any patients with a clinical suspicion of COVID-19 were tested and confirmed COVID-19 negative prior to transfer to the community hospital, as per the policy at that point; guidance was subsequently updated to test all prior to transfer. The patients had significant co-morbidity (mean seven co-morbidities) and polypharmacy (mean eight co-prescribed regular medications). Seven of the patients were asymptomatic for COVID-19 at the point of positive swab. Thirteen of the patients were transferred back to the acute trust, although this did not correlate with severity of symptoms as it was early in the pandemic and these patients were anticipated to potentially require further medical input. Four patients died within 30 days of COVID-19 positive test.

### Serology

Subsequent serological testing was performed in 115 of the 118 staff who were tested by RT-PCR. Twenty-nine staff were PCR positive and serology positive. Ten additional staff members had positive serology. There was one case who was PCR positive and had successful viral sequencing for whom the serology was negative on two repeated tests.

### Viral genome sequence

Viral RNA was available for 28/30 staff and all 16 patients. Near-complete viral genome sequences were successfully generated for 18 staff and 13 patients, plus 122 additional viral genomes from cases in the Exeter area at that point. Median depth of sequencing across each genome ranged between 1,657 x and 27,377 x. Each genome had at least 20 x coverage across 90.16 – 99.87 % of its length.

Two hundred and thirty-one sites in the genome showed single nucleotide polymorphism (SNP). The 31 community hospital samples fell into seven distinct haplotypes (A-H), defined by 29 SNPs that were polymorphic among these; the other 202 SNPs were invariant among the outbreak samples. The relationships between the viral haplotypes are illustrated in Figure 1A, placing the local cases into regional context of viral genetic diversity, and Figure 2. Civet and Pangolin (https://articnetwork.github.io/civet/) assigned all sequenced genomes to global lineage B1.1 and UK lineage UK2266 except for one sample (our haplotype H), which was assigned to global lineage B1 and UK lineage UK5741.

**Figure 1.**
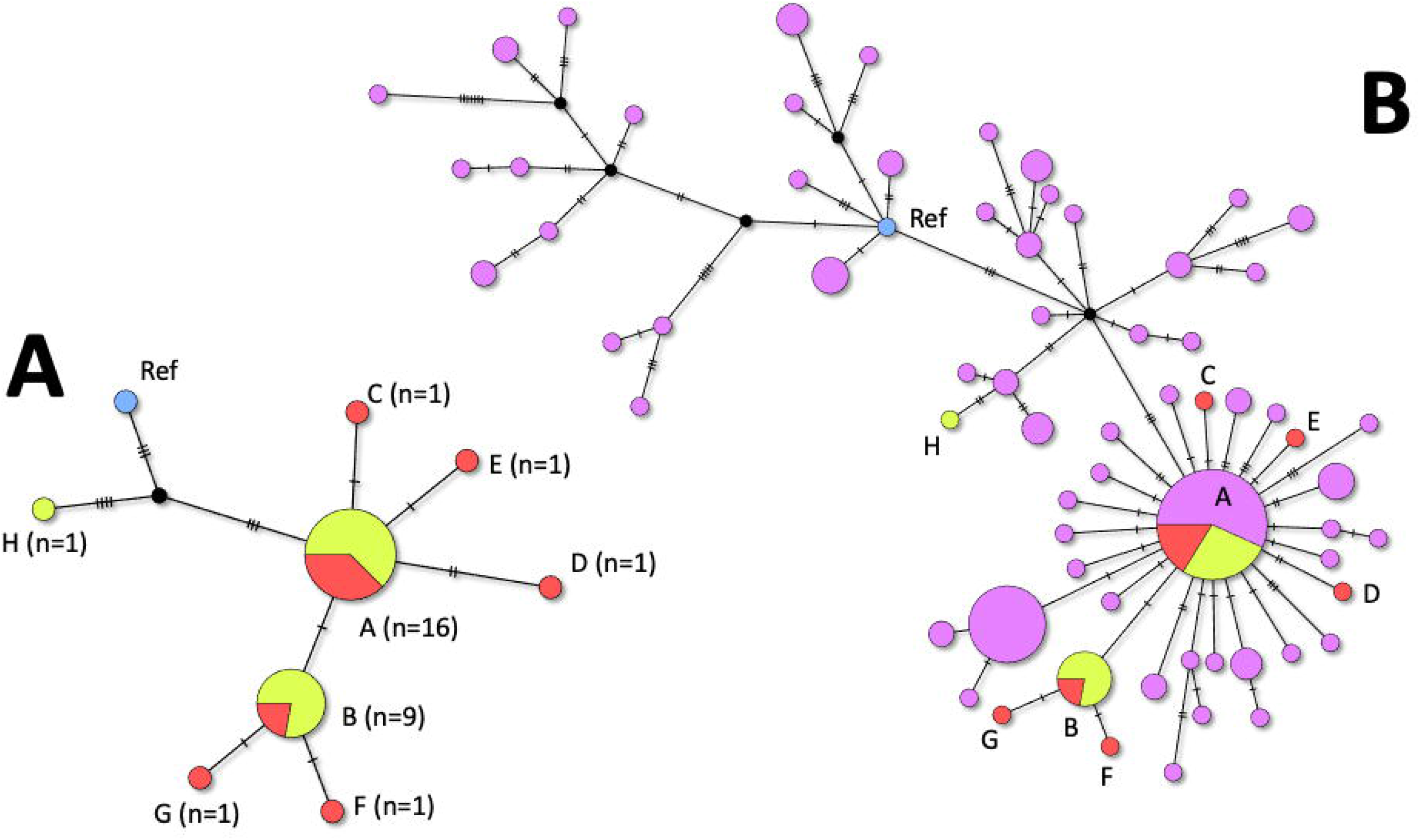
Median-joining networks of the viral haplotypes. Panel A includes only cases from the community hospital outbreak while panel B includes all 141 genomes sequenced at Exeter at that time. Also included in both networks is the Wuhan-Hu-1 reference genome (GenBank: MN908947.3). The genome sequences fell into 61 distinct haplotypes overall (defined by 231 SNPs) and eight distinct haplotypes in the outbreak (defined by 29 SNPs, listed in Supplementary Table 1). Each node represents a single haplotype and its area is proportional to the numbers of samples within that haplotype; the proportions of the circle shaded in red and yellow indicate the proportions of samples in that haplotype from outbreak and non-outbreak samples respectively. The mutations separating the nodes are indicated by the cross hatches on the connecting edges.

**Figure 2:**
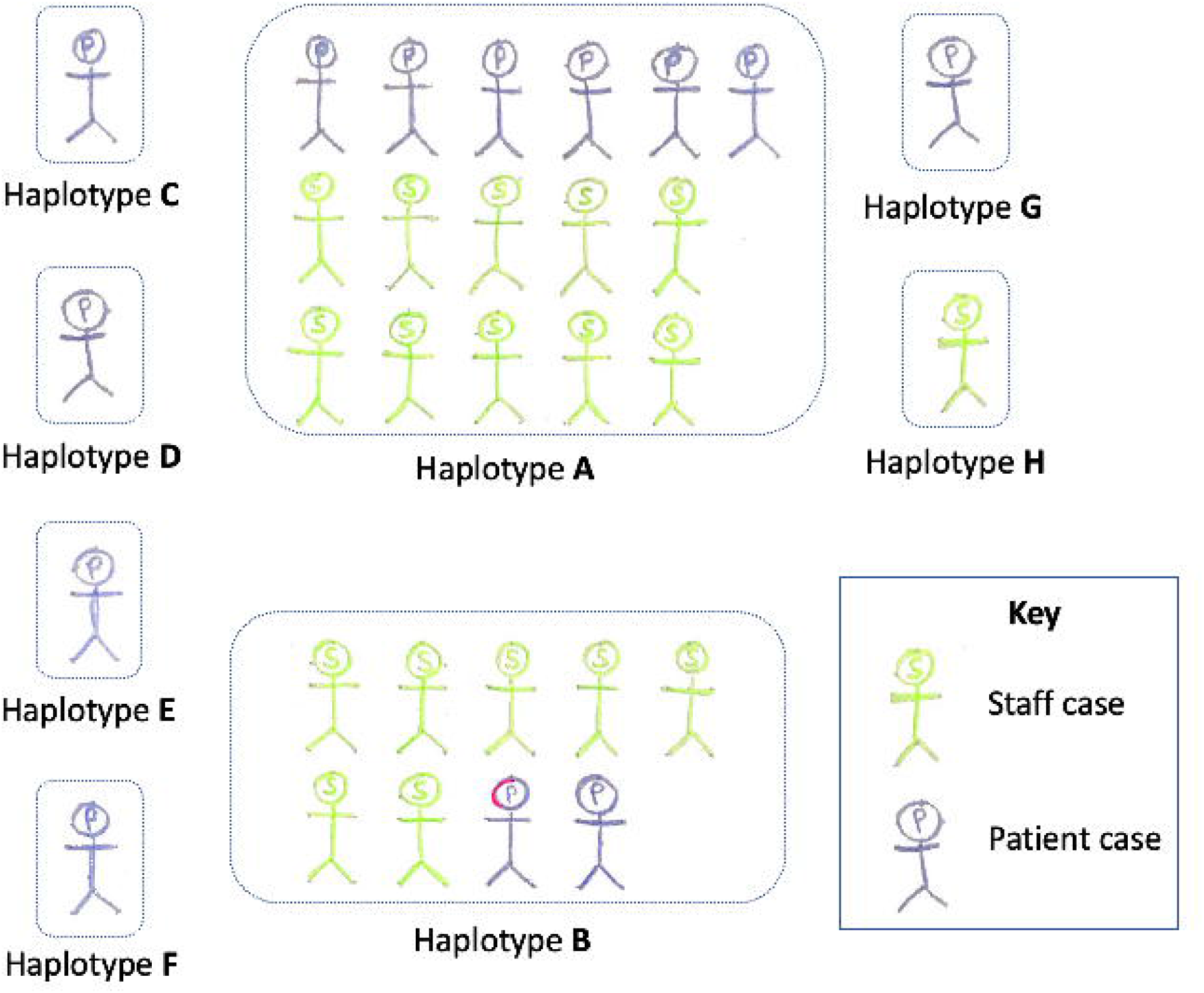
Haplotypes within outbreak with number of cases in patients and staff. The Full details of the haplotypes A – G are listed in Supplementary Table 1.

Overall, the viral sequences from the 13 patients represented seven haplotypes as illustrated in Figure 2. Two were shared with staff: A (n=6 patients) and B (n=2 patients) and five were unique (C, D, E, F and G). The viral sequences from most of the 18 staff fall into two haplotypes A (n=10 staff) and B (n=7 staff). Haplotype H was identified in a new staff member who had not previously had overlapping shifts with other COVID-19 positive staff members. Given the estimated mutation rate of approximately two changes per genome per month^4^, while we cannot confidently propose links between cases whose haplotypes are differentiated by one or two SNPs, it is plausible that they are connected.

The sequencing-based networks in Figure 1 reveal that the most-frequent haplotype (A) in the outbreak cases is frequent across samples sequenced regionally (n= 37 (haplotype A) out of 141 sequenced at that point). This suggests that this haplotype is common and therefore its co-incidence among 16 of 31 cases could be explained by either spread from a single initial case or multiple independent acquisitions from the community, or a combination of both of these. However, the second-most frequent haplotype (B) among outbreak cases is not seen in other samples external to the outbreak (Figure 1), nor in other samples sequenced to date nationally, strongly suggesting that those cases are epidemiologically linked. Two further cases were separated by a single mutation from this haplotype (haplotypes E and F) suggesting they could have arisen from haplotype B, and so were included in analysis of this cluster. We therefore undertook a more detailed analysis on the cluster of cases with haplotype B plus haplotypes E and F to try to identify the likely spread of infection to guide policy to prevent future outbreaks (Figure 3).

**Figure 3:**
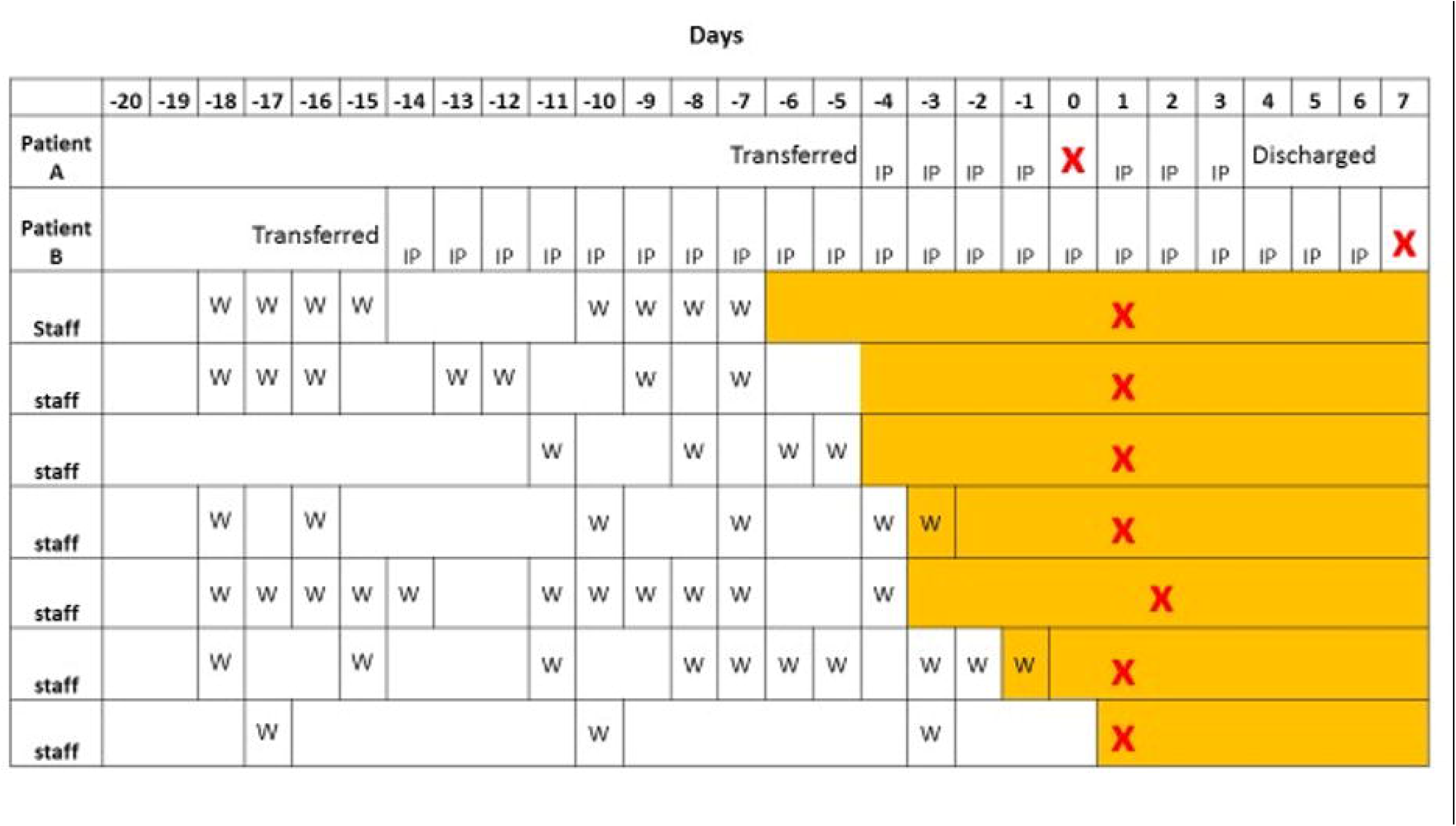
Timeline for haplotype B cases including patient days in community hospital and staff work days. Day 0=first confirmed positive case within cluster. Potential exposure within community hospital is demarcated by ‘W’ for work day in staff and ‘IP’ for inpatient at community hospital for patients. Yellow shading represents symptoms. Positive COVID-19 test is represented by a red cross.

### Cluster analysis

The patients with haplotypes E and F and three of the four patients with haplotype B were asymptomatic and screened for COVID-19 by PCR test as part of the outbreak analysis, with one complaining of a mild cough following positive result. Patient 4 was negative at initial screening and subsequently positive. Detailed case-note review revealed the timeline of exposure and symptom onset (Figure 3). Three staff members within the cluster were symptomatic and off work before any patient cases were confirmed positive and prior to the transfer of patients 1 and 2 from the acute hospital to the community hospital. It is noticeable that the cluster of staff who share haplotype B sequencing had overlapping shift patterns over the two-week period preceding symptom onset, with regular points of contact at shift handover. Additionally, the staff cases within this cluster all developed symptoms within a one-week period.

## Discussion

Genomic sequencing has shown that the majority of staff cases in this outbreak had a limited lineage, while patient cases were more likely to be distinct. The most common haplotype in this outbreak maps to the lineage sequenced most frequently both regionally and nationally. Distinct mutations have arisen in this viral lineage to result in the cluster of cases described with haplotypes B, E and F, which have only been identified in this outbreak.

The cluster of cases within this distinct haplotype suggests a likely transmission pathway within the community hospital. COVID-19 positive patients more frequently had a unique haplotype than did staff, suggesting that patient-to-staff transmission was rare. The staff’s viral genomes predominantly fell within two haplotypes, one of which was unique to this outbreak. At the time of this reported outbreak, the South West of England had the lowest COVID-19 incidence in the country.^18^ Such a large cluster of cases, within two (closely related) haplotypes in staff, indicates it is highly likely that there was significant staff-to-staff transmission in this outbreak. Patients had a more diverse range of haplotypes, but it is also probable that there was some staff-to-patient transmission, given overlapping haplotypes between staff and inpatients who had been in the community hospital for an average of 18 days prior to screening.

Whilst the route of transmission between staff cannot be definitely assigned, it is noted that the staff with shared haplotypes had overlapping shift patterns. Healthcare workers have higher COVID-19 exposure than the general population due to the nature of their work^19^ and have been commonly affected worldwide.^20^ There have been prior reported outbreaks in which healthcare workers have represented the majority of cases affected,^21^ and nosocomial infection rates likely transmitted by healthcare workers have risen, leading to regular healthcare worker testing.^22^ In the UK COVID-19 was found to be RT-PCR positive in 14% of healthcare workers reporting symptoms.^23^ Rivett et al. described large-scale screening of 1,032 asymptomatic healthcare workers, with 3% positive.^24^ Those with symptoms can follow public health measures on isolation, but asymptomatic carriage is problematic.

In this outbreak viral sequencing provided key information on which cases are likely to be linked versus those from different sources. This illustrates the utility of sequencing and supports the use of rapid viral sequencing in the evaluation of COVID-19 outbreaks. We have shown that in future COVID-19 surge episodes or outbreaks whole-genome sequencing could be used effectively to support clinical evaluation. This method could be employed alongside contact tracing to track COVID-19 spread in real-time.

A limitation of the study is that viral sequence was not generated in all cases, due to unavailable samples or insufficient sequence, probably reflecting low viral load at testing. This outbreak was prior to widespread testing among healthcare workers or asymptomatic inpatients outside outbreak scenarios and if events repeated now it is likely that testing would have occurred earlier in individual symptomatic cases.

A key strength of this study is that it combined viral genomic sequencing with epidemiological clinical analysis, RT-PCR and subsequent serological testing to facilitate full outbreak analysis. The screening of all staff members first by RT-PCR and then by serology testing has given a complete picture in this outbreak. The subsequent antibody testing shows that the majority of cases occurred in the staff during this outbreak. The COG-UK genetic sequencing provides excellent coverage for genomic sequencing in the UK and better clinical corroboration could aid understanding in outbreaks and more generally, such as whether new COVID-19 cases over time in the same setting (e.g. a specific care home) are the same haplotype or if distinct viral haplotypes are being introduced by staff or visitors. This could significantly affect policy, but requires centres having access to both clinical detail and viral genomic sequencing, a particular strength of this study, in which the NHS trust and University teams work in close collaboration. It is particularly challenging to combine sequencing and clinical information from non-hospital settings such as care homes due to centralisation of testing practices.

## Conclusion

This case demonstrates the added value of genomic epidemiology in outbreak analysis. The likely staff-to-staff transmission and potential staff-to-patient transmission should be heeded. It reinforces the need for implementing infection control strategies and social distancing within health and social care environments between healthcare workers^19,25^, even in the absence of symptoms. Ward handovers and other meeting events, plus staff rest spaces and office areas may be particular risks and need to be urgently re-evaluated and adapted during the pandemic of this highly infectious virus. The lessons learnt from this are transferrable to hospitals and care home settings, in which staff groups provide care and assistance to older people. Viral genetic sequencing has identified that staff transmission of COVID-19 was important in this community hospital outbreak.

## Supporting information

Table of viral haplotypes

## Data Availability

The sequence data are available under BioProject PRJEB37886 via https://www.ebi.ac.uk/ena/browser/view/PRJEB37886

## Author contributions

Case identification, patient recruitment, data acquisition and clinical interpretation: JM, CA, BK, AC, AH, CB. Process of genetic sequencing and data production: SM, AJ, BT, J W-D, RM, AF, CS, BB, JH, SE, MM. JM, AJ, BT, AH & DS drafted the report. JM and DS are guarantors for the data. All authors approved the final version of the report

## Acknowledgements

We would like to acknowledge the COVID-19 Genomics UK (COG-UK) consortium, which provides the infrastructure for COVID-19 sequencing across the UK. We would like to acknowledge the Royal Devon and Exeter Tissue Bank team, which supported recruitment and data collection via the ExCoSe study.

---

**Supplementary Table 1**

Haplotypes of the sequenced SARS-CoV-2 genomes.

